# Sotrovimab drives SARS-CoV-2 Omicron variant evolution in immunocompromised patients

**DOI:** 10.1101/2022.04.08.22273513

**Authors:** G. Destras, A. Bal, B. Simon, B. Lina, L. Josset

## Abstract

After monoclonal antibody sotrovimab implementation, Rockett et al have warned on March 9th about two resistant mutations in the spike at position 337 and 340 occurring within the first week in four immunocompromised patients infected by a Delta variant and resulting in viable infection up to 25 days. As sotrovimab is currently the only effective treatment against BA.1 lineage of Omicron variant, we investigated the presence of these mutations in our 22,908 Omicron sequences performed from December 2021 to March 2022.

Among 25 Omicron sequences with S:337 and S:340 substitutions, 9 were reported in six patients who had available clinical data and a follow up. All were immunicompromised, and presented a rapid selection of these mutations after sotrovimab monotherapy infusion.

With these findings, we underscore that although these mutations are rare, they have been exclusively reported in immunocompromised patients treated with sotrovimab. We urge to consider monoclonal antibody as monotherapy in immunocompromised patients as a risk for escape mutants selection.

## Manuscript

Sotrovimab is a monoclonal antibody used as monotherapy in outpatients at risk to develop severe COVID-19 disease. Indications include patients with respiratory, cardiac, metabolic, and immunosuppression comorbidities. Rockett and colleagues have recently shown that among 100 patients infected by Delta variant and treated with sotrovimab monotherapy, 4 were immunocompromised and rapidly developed resistant mutations in the spike at positions 337 and/or 340 (S:337 and/or S:340)^1,2^. As sotrovimab is one of the few monoclonal antibodies that retains efficacy against the widely circulating BA.1 sublineage (Omicron variant), monitoring the prevalence of these mutations is crucial^3^. As part of routine genomic surveillance at the National Reference Center from December 2021 to March 2022^4^, we detected S:340 and/or S:337 mutations in 24 out of 18,882 (0 ·13%) and 1 out of 4,025 (0 ·02%) Omicron BA.1 and BA.2 lineages, respectively. These 25 samples corresponded to 18 patients viruses carrying either S:P337 or S:E340 mutations (Table S1). Clinical data were available for 8 patients, all were immunocompromised and had been treated with sotrovimab 0 to 10 days after symptoms onset (Table S2). For 6 patients with a follow-up, S:337 and S:340 mutations were absent before sotrovimab infusion and were detected at low or high relative frequencies (6 to 100%) within a short timeframe (5-18 days). Resistant virus selection was associated with persistent SARS-CoV-2 excretion up to 43 days except for one patient who cleared its infection after convalescent plasma infusion at D24 (Figure1). These results suggest that sotrovimab can rapidly select S:337 and S:340 mutations in BA.1 and BA.2 sublineages (although not effective *in vitro* against BA.2 sublineage^3^). These mutations rarely emerge in the Omicron variant (0 ·03% [2756/10,042,757] of all Omicron sequences reported on the GISAID Database, Table S3). Nonetheless, it is worth noting that they have exclusively been reported after sotrovimab treatment in immunocompromised patients (Rockett et al, and present study). As previously reported for patients treated with bamlanivimab^5^, we urge to consider monoclonal antibody as monotherapy in immunocompromised patients as a risk for escape mutants selection that may hamper viral clearance. Immunocompromised patients treated by monoclonal antibodies should benefit from a reinforced virological follow-up including viral sequencing and viral load.

**Figure 1.**
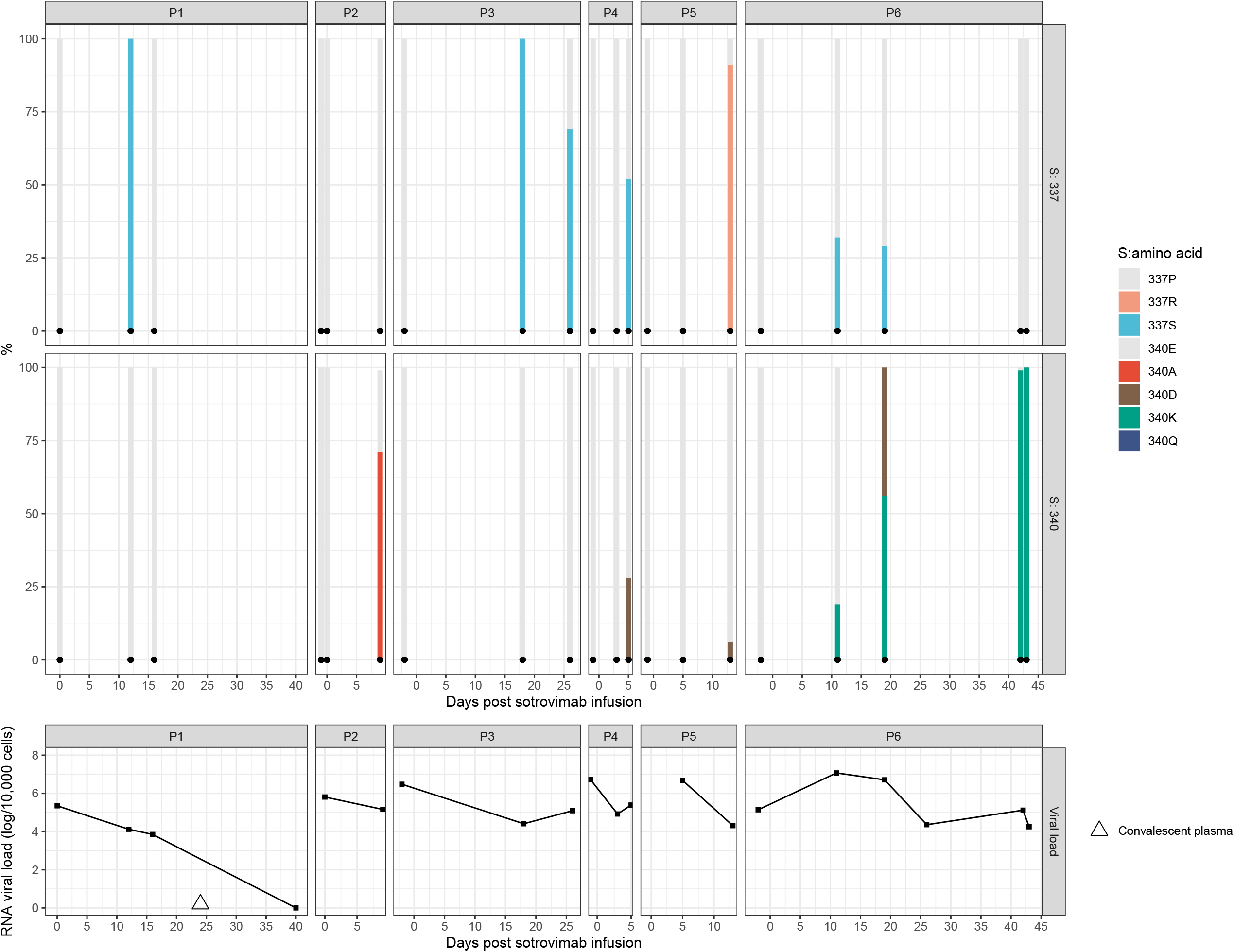
Virological follow-up of immunocompromised patients treated with sotrovimab The above panel shows bars representing relative frequencies of S:337 and S:340 amino-acid substitutions occurring over time after treatment with sotrovimab in immunocompromised patients (n=6). Each colour represents a different amino-acid substitution. Triangle for patient #1 represents convalescent plasma infusion. Normalized SARS-CoV-2viral loads expressed as log10(RNA)/10,000 human cells are represented on the bottom panel.

## Data Availability

All data produced in the present work are contained in the manuscript.

